# Depression and anxiety during the COVID-19 pandemic in Saudi Arabia: a cross-sectional study

**DOI:** 10.1101/2020.05.09.20096677

**Authors:** Hamad S. Alyami, Abdallah Y Naser, Eman Zmaily Dahmash, Mohammed H. Alyami, Musfer S Alyami

## Abstract

**Aims:** The emergence of the COVID-19 global pandemic, with a high transmission and mortality rate, has created an extraordinary crisis worldwide. Such an unusual situation may have an undesirable impact on the mental health of individuals which, in turn, may influence their outcomes. This study aimed to explore the influence of the COVID-19 pandemic on the psychological disposition of residents of the Kingdom of Saudi Arabia.

**Methods:** A cross-sectional study using an online survey was conducted in Saudi Arabia between 27 March and 27 April 2020. The Patient Health Questionnaire (PHQ-9) and Generalised Anxiety Disorder-7 (GAD-7) were used to assess depression and anxiety. Logistic regression analysis was used to identify predictors of these.

**Results:** A total of 2,081 individuals participated in the study. The prevalence of depression and anxiety among the study participants was 9.4% and 7.3%, respectively. Non-Saudi residents, individuals aged 50 years and above, divorced people, retired people, university students, and those with an income between 2,000 and 10,000 SR were at higher risk of developing depression. Saudi individuals, married people, the unemployed, and those with a high income (> 10,000 RS) were at higher risk of developing anxiety.

**Conclusion:** We found that there is a wide range of Saudi residents who are at higher risk of developing mental illness during the current COVID-19 pandemic. Policymakers and mental healthcare providers are advised to provide continuous monitoring of the psychological consequences during this pandemic and provide the required health support.

**What is already known about this subject?:**

– The emergence of the COVID-19 global pandemic, with a high transmission and mortality rate, has created an extraordinary crisis worldwide.
– The COVID-19 pandemic might have an undesirable impact on the mental health of individuals.

**What does this article add?:**

– Depression and anxiety are common among the Saudi population.
– A considerable proportion of the Saudi population is concerned about contracting COVID-19 or transmitting it to family members.
– Unemployed individuals and university students are at higher risk of depression and anxiety.

## 1. Introduction

COVID-19, which stands for coronavirus disease 2019, is a cluster of three acute respiratory illnesses that first occurred in Wuhan, Hubei Province, China in December 2019 ^1^. In early March 2020, the first case of COVID-19 was confirmed in the Kingdom of Saudi Arabia, and since then, it has caused 2,017 deaths out of the 217,108 patients who were infected with this disease until the 7^th^ of July 2020 ^2-4^. The causative agent for COVID-19 has been identified as a new RNA virus from the beta-coronavirus family; its transmission rate is considered high because it is transmitted in different ways, such as respiratory droplets and close contact. The World Health Organization (WHO) has categorised COVID-19 as a pandemic infection since the respiratory illness it causes is highly contagious ^5^ because of the novelty of the virus, its rapid spread, and the lack of therapeutic and preventative strategies ^6^.

The spread of COVID-19 presents serious risks globally and in Saudi Arabia, which has reported 393,377 cases and 6,704 deaths as of 06 April 2021^7^. Saudi Arabia has exceptional circumstances as it is a hub for millions of foreign workers and pilgrims from across the globe. In response to the pandemic and to combat the spread of the disease, the government took swift action and closed the two holy mosques, suspended travel to the country, closed most businesses and limited individuals’ movement. Further, the government created a national narrative to encourage citizens to adhere to the emergency measures established in response to the pandemic^6^. The Kingdom of Saudi Arabia took the deadly coronavirus outbreak seriously, even before the Ministry of Health announced the first confirmed COVID-19 case; for example, it announced the temporary suspension of entry to Makkah and Madinah in February 2020 ^8^. After the first confirmed case, the government announced a series of extreme measures to control the spread of the virus, beginning on 8 March with a ban on all transport in and out of the Qatif Governorate.

Then, on 6 April, they announced a 24-hour curfew to be implemented in the major cities, with movement restricted to essential travel between 6 a.m. and 3 p.m. ^9^ The extremely proactive measures taken to prevent the spread of the virus could have provoked other health outcomes usually neglected in crisis and pandemic management ^10-15^. Recent studies have shown that the rapid global growth in the number of COVID-19 cases and deaths, the collapse of the healthcare system in many countries, and the subsequent lack of effective medical treatment have had the same effect ^16-18^. COVID-19 is having a severe impact on the physical and mental health of the public ^16,18-21^; therefore, we aimed to assess the mental health burden placed on citizens and residents inside the Kingdom of Saudi Arabia at this time, as well as to identify potential populations who may need psychological intervention.

## 2. Methods

### 2.1. Study design and study population

A cross-sectional study using an online survey was conducted in Saudi Arabia between 27 March and 27 April 2020 to explore depression and anxiety among the general population during the COVID-19 pandemic.

### 2.2. Sampling strategy

A convenience sample of eligible participants drawn from the general population was invited through social media (Facebook and WhatsApp) to participate in the study. All the participants participated voluntarily in the study and were, thus, considered exempt from written informed consent. The study aims and objectives were clearly explained at the beginning of the survey.

The inclusion criteria were: a) participants aged 18 years and above and living currently in Saudi Arabia, and b) participants who had no apparent cognitive deficiency. Participants were excluded if they were: a) below 18 years of age, b) unable to understand the Arabic language, and c) unable to participate due to physical or emotional distress. The inclusion and exclusion criteria were clearly stated in the invitation letter accompanying the questionnaire. Participants were invited to participate if they were eligible.

### 2.3. Depression and anxiety assessment scales

Previously validated assessment scales, the Patient Health Questionnaire (PHQ)-9, and Generalised Anxiety Disorder seven-item (GAD-7), were used to assess depression and anxiety among the study participants. These screening instruments have been frequently used and have been validated as brief screening tools among various populations for depression and anxiety ^22-26^. The following information was also collected: the participants’ demographics (age, gender, income, education level, employment status, and marital status). Furthermore, participants were asked whether they were worried about being infected with COVID-19 or transmitting it to family members (yes/no question) and whether they had any underlying chronic conditions (yes/no question).

The PHQ-9 scale is a nine-item instrument given to participants to screen for the presence and severity of depression ^27,28^. The GAD-7 instrument was used to screen for anxiety ^29^. The PHQ-9 and the GAD-7 instruments asked the participants about the degree of applicability of a range of items (questions), using a four-point Likert scale. Participants’ responses ranged from 0 to 3, where zero meant “Not at all” and three meant “Nearly every day”. The PHQ-9 instrument includes nine items. Items are scored from 0 to 3, generating a total score ranging from 0 to 27. A total score of 0–4 indicates minimal depression, 5–9 mild depression, 10–14 moderate depression, 15-19 moderately severe depression, and 20–27 severe depression ^30^. The GAD-7 instrument includes seven items. Items are scored from 0 to 3, generating a total score ranging from 0 to 21. A total score of 5–9 indicates mild anxiety, 10–14 moderate anxiety, and 15–21 severe anxiety ^31^.

### 2.4. An estimate of prevalence and classification of depression and anxiety

Prevalence rates of depression and anxiety were determined using a cut-off point, as recommended by the authors of the PHQ-9 and GAD-7 scales. In this study, depression was defined as a total score of (≥ 15) in the PHQ-9 instrument, indicating a case with moderately severe or severe depression. Anxiety was defined using the GAD-7 instrument with a total score of (≥ 15), indicating a case of severe anxiety. The higher the score, the more severe the case identified by either scale.

The prevalence rate of depression was estimated by dividing the number of participants who exceeded the borderline score (≥ 15) by the total number of participants in the same population. The prevalence rate of anxiety was calculated using the same procedure.

### 2.5. Sample size

The target sample size was estimated based on WHO recommendations for the minimal sample size needed for a prevalence study ^32^. Using a confidence interval of 95%, a standard deviation of 0.5, a margin of error of 5%, the required sample size was 385 participants.

### 2.6. Statistical analysis

Descriptive statistics were used to describe the participants’ demographic characteristics. Continuous data were reported as mean ± SD for normally distributed variables and median (interquartile range (IQR)) for non-normally distributed variables. Categorical data were reported as percentages (frequencies). The Mann-Whitney U test/Kruskal-Wallis test was used to compare the median scores between different demographic groups. Logistic regression was used to estimate the odds ratios (ORs) with 95% confidence intervals (CIs) for anxiety or depression. Logistic regression models were carried out using anxiety (≥ 15) or depression scores (≥ 15) above the cut-off points highlighted above. A two-sided p<0.05 was considered to be statistically significant. The statistical analyses were carried out using SPSS (version 25).

### 2.7. Ethical considerations

Ethical approval was obtained from the School of Life and Health Sciences at Najran University, Najran, Saudi Arabia (27/03/2020ET). As participation in the study was voluntary and the study was on the general public without any intervention, meaning that it involved no more than minimal risk, the Research Ethics Committee approved a consent waiver.

## 3. Results

### 3.1. Participant characteristics

A total of 2,081 individuals participated in the study. **Table 1** details their baseline characteristics. The majority were Saudi (n= 1,765, 84.8%), (n=1,404, 67.5%) males, aged between 30 - 49 years (n= 1,140, 54.8%), married (n= 1,298, 62.4%), holding a bachelor’s degree (n= 1,284, 61.7%). More than half of them were employed (n= 1,272, 61.1%). Around 12.3% (n= 255) of the participants reported that they had a history of chronic disease. The majority (n= 1,587, 76.3%) of them reported that they were concerned about contracting COVID-19 or transmitting it to family members. When they were asked whether they had identified any problems over the previous two weeks, and the extent to which these problems had prevented them from doing their work, looking after their household affairs or dealing with people, around half of them (n= 1,053, 50.6%) reported that they faced difficulties.

**Table 1.** Participants baseline characteristics

| **Demographics** | **N. (%)** |
| --- | --- |
| **Nationality No. (%)** |  |
| Saudi | 1,765 (84.8) |
| **Gender No. (%)** |  |
| Male | 1,404 (67.5) |
| **Age No. (%)** |  |
| 18 – 29 years | 757 (36.4) |
| 30 – 49 years | 1,140 (54.8) |
| 50 years and above | 184 (8.8) |
| **Marital Status No. (%)** |  |
| Single | 652 (31.3) |
| Married | 1,298 (62.4) |
| Divorced | 131 (6.3) |
| **Education level No. (%)** |  |
| Completed secondary grade | 488 (23.5) |
| Complete bachelor degree | 1,284 (61.7) |
| Higher education | 309 (14.8) |
| **Employment status No. (%)** |  |
| Retired | 79 (3.8) |
| Unemployed | 455 (21.9) |
| Employed | 1,272 (61.1) |
| University students | 275 (13.2) |
| **Income No. (%)** |  |
| 2000 SR or below | 585 (28.1) |
| 2000 – 5000 SR | 273 (13.1) |
| 5000 – 10,000 SR | 614 (29.5) |
| 10,000 SR and above | 609 (29.3) |
| **Chronic disease history No. (%)** |  |
| Yes | 255 (12.3) |
| **Worried about being infected with the corona virus or transmitting it to family members No. (%)** |  |
| Yes | 1,587 (76.3) |

No: Number, SR: Saudi Riyal

### 3.2. Prevalence of mental health problems

The prevalence of depression among the participants was 9.4% (n= 196). The proportions of minimal, mild, moderate, moderately severe, and severe depression were 29.6%, 40.9%, 20 %, 6.2%, and 3.2%, respectively. The prevalence of anxiety among the participants was 7.3% (n= 151). The proportions of mild, moderate, and severe anxiety were 73.5%, 19.3%, and 7.3%, respectively. **Table 2 and Table 3** detail the prevalence of depression and anxiety among the participants, stratified by severity and gender, respectively.

**Table 2.** Prevalence of depression and anxiety among the participants stratified by severity

|  | N. (%) |
| --- | --- |
| **Depression diagnose** |  |
| **Minimal depression** | 617 (29.6) |
| **Mild depression** | 852 (40.9) |
| **Moderate depression** | 416 (20.0) |
| **Moderately severe depression** | 129 (6.2) |
| **Severe depression** | 67 (3.2) |
| **Anxiety diagnose** |  |
| **Mild anxiety** | 1,529 (73.5) |
| **Moderate anxiety** | 401 (19.3) |
| **Severe anxiety** | 151(7.3) |

**Table 3.** Prevalence of depression and anxiety among the participants stratified by severity and gender

|  | N. (%) |  |
| --- | --- | --- |
| Depression diagnose |  |  |
|  | Males | Females |
| Minimal depression | 455 (32.4) | 162 (23.9) |
| Mild depression | 565 (40.2) | 287 (42.4) |
| Moderate depression | 270 (19.2) | 146 (21.6) |
| Moderately severe depression | 75 (5.3) | 54 (8.0) |
| Severe depression | 39 (2.8) | 28 (4.1) |
| Anxiety diagnose |  |  |
|  | Males | Females |
| Mild anxiety | 1,072 (76.4) | 457(67.5) |
| Moderate anxiety | 259 (18.4) | 142 (21.0) |
| Severe anxiety | 79 (5.6) | 72 (10.6) |

### 3.3. Participant demographics and mental health problems

**Table 4** presents the participant demographics and their median depression and anxiety scores. The depression median score significantly differed across participants with different demographical characteristics (p<0.01). The anxiety median score significantly differed across participants by nationality, gender, and education level (p<0.05).

**Table 4.** Depression and anxiety median score stratified by participants’ characteristics

|  | Depression score |  |  | Anxiety score |  |  |
| --- | --- | --- | --- | --- | --- | --- |
| Variable | Median | IQR | P-value | Median | IQR | P-value |
| Nationality |  |  |  |  |  |  |
| Saudi | 7.00 | 6.00 | 0.000*** | 6.00 | 5.00 | 0.000*** |
| Non-Saudi | 9.00 | 7.00 |  | 7.00 | 5.00 |  |
| Gender |  |  |  |  |  |  |
| Males | 7.00 | 7.00 | 0.000*** | 5.00 | 5.00 | 0.000*** |
| Females | 8.00 | 6.00 |  | 6.00 | 5.00 |  |
| Age |  |  |  |  |  |  |
| 18 –29 years | 8.00 | 7.00 | 0.001** | 5.00 | 6.00 | 0.659 |
| 30 – 49 years | 7.00 | 6.00 |  | 6.00 | 4.00 |  |
| 50 years and above | 8.00 | 8.00 |  | 6.00 | 4.00 |  |
| Marital status |  |  |  |  |  |  |
| Single | 8.00 | 8.00 | 0.000*** | 5.00 | 6.00 | 0.228 |
| Married | 7.00 | 5.00 |  | 6.00 | 5.00 |  |
| Divorced | 8.00 | 6.00 |  | 6.00 | 4.00 |  |
| Education level |  |  |  |  |  |  |
| Completed secondary grade | 8.00 | 8.00 | 0.005** | 6.00 | 7.00 | 0.012* |
| Complete bachelor degree | 7.00 | 6.00 |  | 6.00 | 5.00 |  |
| Higher education | 7.00 | 6.00 |  | 6.00 | 4.00 |  |
| Employment status |  |  |  |  |  |  |
| Retired | 7.00 | 8.00 | 0.000*** | 6.00 | 6.00 | 0.188 |
| Unemployed | 7.00 | 7.00 |  | 6.00 | 5.00 |  |
| Employed | 7.00 | 6.00 |  | 6.00 | 5.00 |  |
| University students | 8.00 | 8.00 |  | 5.00 | 5.00 |  |
| Income |  |  |  |  |  |  |
| 2000 SR or below | 8.00 | 8.00 | 0.000*** | 6.00 | 6.00 | 0.701 |
| 2000 – 5000 SR | 7.00 | 7.00 |  | 6.00 | 4.00 |  |
| 5000 – 10,000 SR | 7.00 | 6.00 |  | 6.00 | 4.00 |  |
| 10,000 SR and above | 6.00 | 6.00 |  | 6.00 | 5.00 |  |

\*p < 0.05; \*\*p < 0.01; \*\*\*p < 0.001; Abbreviation: IQR, Interquartile Range

Non-Saudi residents, females, elderly individuals aged 50 years and above and young individuals aged below 29 years, divorced and single people, individuals with a low education level, university students, and individuals with a low income (2000 SR and below) tended to have higher depression median scores compared to the others. Non-Saudi residents, females, and individuals with low education levels tended to have higher anxiety median scores compared to the others.

The logistic regression analysis identified the following groups to be at a higher risk of depression: a) unemployed individuals and b) university students. Meanwhile, the following groups were at a lower risk of depression: a) Saudi residents, b) males, c) married individuals, d) individuals who had completed a bachelor’s degree, and e) individuals with a high income (5,000 SR and above). Furthermore, logistic regression analysis showed that the following groups were at a higher risk of anxiety: a) unemployed individuals, and b) and university students. On the other hand, the following groups were at a lower risk of anxiety: a) males, b) elderly individuals (aged 50 years and above), c) divorced individuals, d) individuals with moderate income (5,000 SR to 10,000 SR). See **Table 5**.

**Table 5.**
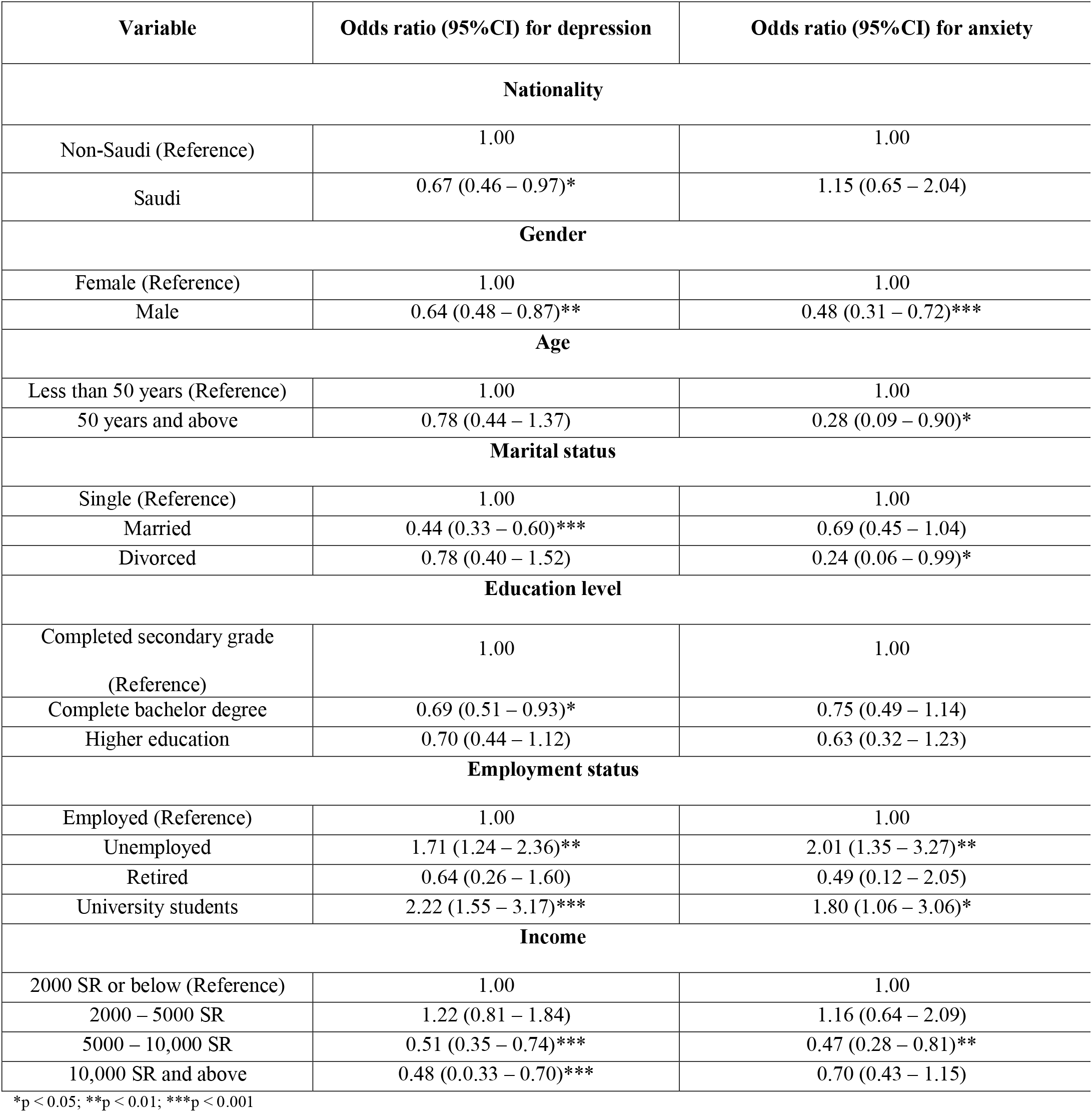
Logistic regression analysis

## 4. Discussion

Overall, the findings in this study demonstrated that more than 9.4% of respondents had moderate to severe depression, whereas the prevalence of moderate to severe anxiety exceeded 7.3%. A previous systematic review that explored the impact of COVID-19 on mental health in the general population reported a much higher rate of symptoms of anxiety (ranged between 6.3% to 50.9%) and depression (14.6% to 48.3%) in different countries such as China, Spain, Italy, Iran, the US, Turkey, Nepal and Denmark ^21^. Some of these countries have reported comparable rates to our results. Another recent study in Vietnam reported a close rate of anxiety and depression, with respect to our estimates, with 4.9% and 7.0%, respectively^19^. On the other hand, a previous study in the Philippines has reported higher prevalence rates of moderate to severe depression (16.9%) and anxiety (28.8%) ^20^. There are many reasons for the wide range of estimates across different countries. One of these could be different assessment tools used. The timing of the study is another important influencing factor as the psychological impact was expected to be much more intense at the beginning of the pandemic since little information was known about the disease. This could have provoked public fear and other psychological symptoms. During stressful conditions, as is the case with the COVID-19 pandemic, fear and anxiety about the disease can be so overwhelming that it may cause depression and anxiety among both adults and children ^33^. The sudden shutdown of services and lockdown of people makes them vulnerable, particularly when dealing with the unpredictable status of the outbreak. The fear of getting the disease and losing loved ones is another predisposing factor that may result in such a condition ^34^. The prevalence rate of moderate to severe depression symptoms in this study seems to be considerably higher than the one reported by the Chinese study, which included 1,210 respondents, during the COVID-19 outbreak (16.5%), whereas comparable rates for anxiety were recorded (28.8%) ^3^. Also, it has been observed that results vary according to the sample size and the assessment tool used. Another nationwide study among Chinese people during the pandemic, which included 52,730 participants, revealed a psychological distress prevalence rate of 35% among all respondents. Distress symptoms, according to the employed assessment tool, included depression and anxiety ^35^.

This study also revealed that non-Saudi residents had significantly higher prevalence rates of depression and anxiety symptoms (p< 0.001) than Saudi individuals. Such results aligned with the percentage of infected cases among the two groups, where the data showed a pattern in terms of who was more likely to become infected with the virus. Recent reports have shown that, among confirmed cases, Saudis accounted for 19% of total cases, while other nationalities accounted for 81%. These results could be attributed to the status of most non-Saudis in terms of occupation and residence status. A substantial number of foreign workers are in the labour force and live in heavily crowded areas where they are often unable to observe social distancing requirements ^36^. Previous studies have reported that foreign workers experienced the highest level of distress among all occupations. Reasons such as worrying about exposure to the virus in public transportation when commuting to and from work, delays in work time, and job security and the subsequent loss of their salary may explain the high stress levels ^35^. Such results oblige the government to take and reinforce specific measures to control the increase of infected cases.

The American Department of Labor has provided a list of recommendations to reduce the possibility of worker infection during the pandemic. These include wearing cloth face coverings, at a minimum, at all times when around co-workers or the general public, frequently washing your hands with soap and water for at least 20 seconds, avoiding touching your eyes, nose or mouth with unwashed hands, practicing good respiratory etiquette, including covering coughs and sneezes or coughing/sneezing into your elbow/upper sleeve, avoiding close contact (within 6 feet for a total of 15 minutes or more over a 24-hour period) with people who are visibly sick and practicing physical distancing with co-workers and the public, staying home if sick, and recognizing personal risk factors ^37^.

Depression and anxiety symptoms were more likely to occur in women than men. Such results agree with other studies that have investigated depression and anxiety among the Saudi Arabian population. The results of a study conducted by Al-Khathami et al. (2002) reported that the prevalence of minor mental illness was significantly higher in women (22.2%) than men (13.7%) (p =0.0073) ^38^. This confirms what has been reported in a previous meta-analysis that explored the prevalence of depression across 30 countries all over the world between 1994 and 2014. In this study, women showed higher depression rates compared to men: 14.4% (95% CI: 11.1% to 11.7%) for women and 11.5% (95% CI: 9% to 14.6%) for men ^39^.

Further, the prevalence rate was higher in the younger age group, which agrees with our study, as a higher score of depression was associated with individuals younger than 29 years. Further, the study of Wang et al. (2020) revealed that women were significantly associated with a greater psychological impact following the COVID-19 outbreak and had higher levels of stress, anxiety and depression (p < 0.05)^1^. Several factors could have contributed to the women’s higher depression and anxiety prevalence rates, including biological sex differences, culture, diet, female hormonal fluctuations, or education ^40^.

Sociodemographic variables associated with depression and anxiety were assessed using logistic regression analysis. The results showed that individuals over the age of 50 suffered from higher depressive symptoms, as do those who are single or with lower education levels. Similar findings were reported by Wang et al. (2020), suggesting an association of lower education with a greater likelihood of depression during the pandemic. Further, our findings provided data that suggested the levels of anxiety and depression-related symptoms were greater among students and the unemployed, or those with a low income. The results agreed with previous research, which also found that students were more likely to have depression and anxiety ^3^. The onset of the pandemic was in the middle of the academic year, which may have prompted the students’ fear of losing the year or the occurrence of delays in their studies This was in addition to their lack of confidence in distance learning. The lockdown and social distancing are expected to remain in place for some time to come, and this will have a direct effect on low-income and unemployed individuals ^41^; this might put such categories under a higher level of stress, which could lead to symptoms of anxiety and/or depression.

The results of this research have emphasised the impact of the COVID-19 pandemic on the mental health of individuals, expressed in depression and anxiety. Many aspects of the findings agree with those reported during the pandemic in other countries. Therefore, a worldwide collaborative effort is required to develop measures that can address mental health during such pandemics and manage it. The most evidence-based treatment is cognitive behaviour therapy (CBT), especially Internet CBT as it can prevent the spread of infection during the pandemic. CBT could be used to treat psychiatric symptoms during the current situation ^42^. It provides a cost-effective option that could alleviate negative psychological impacts ^43^. A previous meta-analysis has proved the effectiveness of internet CBT in treating psychiatric symptoms such as insomnia ^44^.

This study has demonstrated several strengths. First, it has addressed the prevalence of depression and anxiety within the initial phase of the pandemic and, hence, may provide valuable information to policymakers that will enable them to make informed decisions and introduce psychological interventions that can minimise untoward psychological effects on the mental health status of the Saudi Arabian population. Second, the study has employed validated tools for the assessments which have enhanced the reliability of the study. Third, it involved an acceptable sample size that was not limited to specific geographical areas of Saudi Arabia. However, this study has several limitations. First, the study was based on a web-based survey method, so some vulnerable individuals who have no access to the internet and are unfamiliar with online questionnaires were missed. Second, due to the sudden occurrence of the outbreak, an individual’s anxiety and depression prevalence before the outbreak could not be gauged. Third, the survey was administered at a single period and so the stability of the responses is unknown. The study design itself, a cross-sectional survey design, limited our ability to identify causality between study variables, unlike a recent longitudinal study that was conducted in China which was able to monitor the change in psychological status among the general population across different time points of the pandemic ^45^. Fourth, the sample may be biased as those who were more interested in mental health or distressed by the pandemic may have been more likely to participate. Finally, this study mainly used self-reported questionnaires to measure psychiatric symptoms and did not make clinical diagnoses. The gold standard for establishing psychiatric diagnosis involves structured clinical interviews and functional neuroimaging ^46,47^.

## 5. Conclusion

During the initial phase of the COVID-19 pandemic in Saudi Arabia, more than 29% of the respondents had moderate to severe depression, and 26.6% reported moderate to severe anxiety. Female gender, student status, low income and low education level respondents were associated with a greater psychological impact of the outbreak, and higher levels of anxiety and depression. Our findings may enable policymakers to introduce several measures and psychological interventions that can enhance mental health during the current pandemic.

## Data Availability

The data that support the findings of this study are available from the corresponding author upon reasonable request.

## Acknowledgment

This paper appears on a pre-print server (medrxiv): https://www.medrxiv.org/content/10.1101/2020.05.09.20096677v1

## Data Availability Statement

Data sharing is not applicable to this article as no new data were created or analyzed in this study.

